# Thalamic transcranial electrical stimulation with temporal interference enhances sleep spindle activity during a daytime nap

**DOI:** 10.64898/2026.02.20.26346398

**Authors:** Simone Bruno, Beril Mat, Erin L. Schaeffer, Ido Haber, Zhiwei Fan, Sean P. Prahl, Mackenzie R. Wilcox, E. Strainis, Madelynn D. Loring, Tariq Alauddin, Richard F. Smith, Peter Achermann, Stefan Beerli, Myles Capstick, Esra Neufeld, Niels Kuster, William Marshall, Larissa Albantakis, Stephanie G. Jones, Chiara Cirelli, Melanie Boly, Giulio Tononi

**Affiliations:** Department of Psychiatry, University of Wisconsin-Madison, Madison, Wisconsin; Medical Scientist Training Program, University of Wisconsin-Madison, Madison, Wisconsin; Department of Biomedical Engineering, University of Wisconsin-Madison, Madison, Wisconsin; International Institute for Integrative Sleep Medicine (WPI-IIIS), Tsukuba Institute for Advanced Research (TIAR), University of Tsukuba, Tsukuba, Japan; Institute of Pharmacology and Toxicology, University of Zurich, Zurich, Switzerland; Foundation for Research on Information Technologies in Society (IT’IS), Zurich, Switzerland; Department of Information Technology and Electrical Engineering, Swiss Federal Institute of Technology (ETH) Zurich, Zurich, Switzerland; Department of Mathematics and Statistics, Brock University, St. Catharines, Ontario, Canada; Wisconsin Institute for Sleep and Consciousness, University of Wisconsin-Madison, Madison, Wisconsin; Department of Neurology, University of Wisconsin-Madison, Madison, Wisconsin

**Keywords:** Sleep spindles, Sleep, Non-REM, Thalamus, Thalamo-cortical network, Temporal interference stimulation, Transcranial electrical stimulation, Non-invasive brain stimulation, High-density electroencephalography

## Abstract

**Introduction:** Sleep spindles are electroencephalographic elements characteristic of non-rapid eye movement sleep generated by thalamo-cortical interactions. Spindles have been linked to some of the cognitive benefits afforded by sleep and high spindle activity is associated with increased arousal threshold. Here, we demonstrate that targeting the thalamus with Transcranial Electrical Stimulation with Temporal Interference (TES-TI) can enhance spindle activity.

**Methods:** 46 participants (24 ± 9.5 years; 58.7% F) underwent thalamic TES-TI stimulation during daytime naps. Three stimulation protocols with 15kHz carrier frequency were tested during stage 2 of non-rapid eye movement sleep (N2): fixed difference frequency of 10 Hz (TES^15k^^Hz^-TI^10Hz^), difference frequency matched to individual spindle peak (TES^15k^^Hz^-TI^Peak^), and no difference frequency (TES^15k^^Hz^). Spectral power in the spindle (sigma) band and integrated spindle activity (ISA) were compared before and during the stimulation, and across stimulation protocols.

**Results:** TES^15k^^Hz^-TI^10Hz^ stimulation was associated with a significant increase in sigma band power (Δx̃_STIM-PRE_ = 0.46 log_10_µV^2^, p = 0.0042) and ISA (Δx̃_STIM-PRE_ = 4.064 µV/s, p = 0.030). Cluster-based analysis localized the increase in sigma power across the entire scalp (p = 0.008). Linear mixed effects models showed that both sigma band power and ISA during stimulation increased significantly more in TES^15k^^Hz^-TI^10Hz^ compared to the other experimental condition.

**Conclusions:** This study provides evidence supporting the successful use of TES-TI targeting the thalamus to enhance sleep spindle activity. Stimulation at a fixed difference frequency of 10 Hz increased sigma band power and ISA, whereas neither stimulation matched to individual sigma band peak nor TES alone produced comparable effects. These promising results warrant further investigations into the cognitive and clinical impact of TES-TI, a non-invasive neuromodulation tool that can reach deep brain regions.

**Statement of significance:** This study provides evidence that thalamo-cortical networks, which are central to many physiological and pathological brain activities, can be modulated non-invasively in humans. More specifically, the findings show that transcranial electrical stimulation with temporal interference targeting the thalamus can selectively enhance sleep spindle activity. This work introduces a new strategy for precisely targeting sleep-generating mechanisms regulated by deep brain circuits without surgery or medication. Key next steps include determining whether this increase in spindle activity can positively impact cognition and assessing the translational potential of this approach for clinical populations.

## Introduction

Sleep spindles are transient, burst-like electroencephalographic (EEG) features that serve as a defining characteristic of non-rapid eye movement (NREM) sleep in mammals. First identified by Loomis and colleagues^1^, they were named for their distinctive waxing-and-waning morphology. In human adults, sleep spindles typically show oscillations within the 11–16 Hz frequency range, representing the most important contributor to the EEG sigma band power. Slow (< 13 Hz) and fast (>= 13 Hz) spindles are preferentially found in the frontal and centro-parietal neocortex, respectively^2,3^. Sleep spindles first emerge at the onset of stage N2 sleep and continue to occur into stage N3, where their density and visual detectability decline^4^. Evidence from animal model studies has also demonstrated a role for sleep spindles in guiding the transition from NREM to REM sleep via a transitional stage, which shows greater sigma band power compared with both NREM and REM sleep^5,6^. At the cellular level, spindle activity originates in the thalamus. Specifically, the thalamic reticular nucleus represents the spindle pacemaker and its rhythmic interactions with the thalamocortical relay neurons constitute the core neurophysiological mechanism underlying spindle generation^2,7^.

Growing evidence indicates that sleep spindles, alone or in coordination with other sleep-related oscillations^8^, support sensory gating^2^ and memory consolidation^9^. Sleep spindles deficits have been reported in patients affected by schizophrenia^10^, Alzheimer disease^11^, Parkinson disease^12^ and epilepsy^13^, in association with decreased cognitive abilities, thus representing a potential biomarker and treatment target^14^.

Efforts to enhance sleep spindle activity have employed both pharmacological^15^ and non-invasive brain stimulation approaches^16^. Among central nervous system-active compounds, benzodiazepines and Z-drugs have demonstrated the most robust effects in increasing spindle activity^15^, with associated improvements in memory consolidation^17^. However, their clinical utility is limited by adverse effects, including dependence, cognitive impairment, and suppression of slow wave sleep^18^. Acoustic stimulation in the sigma frequency range has also been explored, though results have been inconsistent^19,20^. Moreover, auditory stimulation presents practical challenges, as the thalamo-cortical system is highly sensitive to acoustic stimuli during sleep^21^, which may elicit unintended awakenings. Transcranial Electric Stimulation (TES), specifically Transcranial Alternating Current Stimulation (tACS), when applied bifrontally at 12 Hz in phase with endogenous spindles, can enhance post-stimulation spindle activity^22^. Despite this, tACS is suboptimal for spindle modulation due to its limited ability to target deep brain structures such as the thalamus without causing significant discomfort to the participant. Additionally, the artifact introduced by conventional TES precludes the acquisition of EEG data concurrent with the stimulation.

Transcranial Electrical Stimulation with Temporal Interference (TES-TI) can address the limitations inherent in conventional non-invasive brain stimulation techniques. This approach employs the use of two high-frequency signals, in the range of kHz, with slightly different frequencies (carriers). Their superposition produces an interference pattern characterized by amplitude modulation of the electric field at the difference frequency^23^. The modulation magnitude is maximal where the currents overlap, which can be used to target neural tissue within a region of interest (ROI) deep in the brain with higher focality compared to standard TES, which always exposes overlaying cortical regions more strongly^24^. Moreover, the use of kHz carriers can virtually eliminate local sensation and discomfort^25,26^, and therefore the risk of awakening subjects during sleep.

In this study, we show how TES-TI can be effectively applied to target the thalamus without disrupting sleep. Through concurrent high-density EEG (hd-EEG) recordings enabled by custom hardware, we show that TES-TI produces a significant enhancement of sleep spindle activity.

## Materials and Methods

### Experimental design

Data for this study were collected during the Rapid Eye Movement Restoration and Enhancement of Sleep-derived Trauma-adaptation (REM REST) clinical trial (ClinicalTrials.gov, NCT06547086), a single-blind longitudinal study aimed at identifying the most effective TES-TI stimulation parameters for eliciting sleep spindles and sawtooth waves during daytime sleep episodes, with the ultimate goal of enhancing emotional well-being and stress resilience.

During the initial (baseline) visit to the Wisconsin Sleep Center, participants were fitted with an hd-EEG net and given the opportunity to take a 90-minute nap. EEG data from the baseline session were used to determine each participant’s individual sigma peak frequency, defined as the 0.25 Hz frequency bin with the highest power within the 11–16 Hz range.

Participants who successfully slept during the baseline nap underwent a Magnetic Resonance Imaging (MRI) session, during which T1- and T2-weighted structural images were acquired. These images were used to optimize electrode placement for TES-TI stimulation, maximizing both intensity and focality of thalamic targeting. Stimulating electrodes were embedded in the hd-EEG net replacing recording electrodes.

In two subsequent visits, participants received the following stimulation protocols during NREM sleep, presented in random order:

- TES-TI, with a 10 Hz difference frequency (TES^15k^^Hz^-TI^10Hz^)
- TES-TI, with a difference frequency matched to the individual sigma peak (TES^15k^^Hz^-TI^Peak^)
- TES, at the carrier frequency of 15 kHz (TES^15k^^Hz^)

All visits were scheduled such that each consecutive nap occurred at least one week after the previous one. Each stimulation interval lasted 180 s, and at least 6 minutes were allowed between two consecutive protocols. Only stimulation intervals entirely included in sleep stage N2, as identified during offline sleep staging, were included in the analysis. Intervals were required to be at least 90 seconds in duration after artifact removal and to be preceded by a minimum of 3 minutes of continuous N2 sleep. Post-stimulation intervals were not included in the present analysis. Indeed, for approximately 50% of the stimulation intervals delivered during N2 sleep, the stimulation period was followed by a transition to a different sleep stage. The reduced number of stimulation intervals followed by the N2 sleep stage made it difficult to distinguish post-stimulation effects from changes attributable to the change in sleep stage. Additional stimulations, whose results are not included in the present paper, were applied during REM sleep and wakefulness using different stimulation parameters and/or targets, in random order across different study visits.

All participants provided written informed consent prior to participation. The study was conducted in accordance with the Helsinki Declaration and approved by the Institutional Review Board of the University of Wisconsin–Madison on 5/28/2024 (ID: 2024-0352).

### Study participants

Eligible participants were English-speaking, medically healthy adults aged 18–50 years who reported habitually taking more than one nap per week and had no history of neurological or sleep disorders. Full eligibility criteria can be found on the ClinicalTrials.gov website.

### EEG data acquisition and preprocessing

EEG data were collected using the Magstim Electrical Geodesics Inc. (EGI) system equipped with 256 silver-silver chloride electrodes, referenced to Cz. Data acquisition and real-time monitoring were performed using EGI Net Station software (v.5.4.2), which enabled experimenters to visualize EEG activity and initiate stimulation based on sleep stage. For sleep staging purpose, the EEG montage included additional electrodes for polysomnographic recordings: electrooculography, recorded using two electrodes placed adjacent to the outer canthi, each referenced to the contralateral mastoid; electromyography, recorded using a pair of electrodes positioned in the submental region; electrocardiography, recorded using two electrodes placed below the clavicle and on the contralateral side of the rib cage. An optical trigger box was connected to both the TES-TI stimulator and the EGI amplifier, marking the onset and offset of stimulation periods to enable precise offline identification of stimulation intervals. All signals were sampled at 500 Hz.

Sleep staging was performed by a registered sleep technician in accordance with the most recent guidelines of the American Academy of Sleep Medicine (AASM)^4^, using the open-source MATLAB-based software CountingSheepPSG^27^.

EEG data preprocessing was conducted using a custom MATLAB routine that incorporated functions from the EEGLAB toolbox^28^. EEG data were bandpass filtered between 0.5 and 40 Hz applying a zero-phase, forward-and-reverse finite impulse response filter using the EEGLAB’s *pop_eegfiltnew* function. Bad channels were identified through visual inspection of the EEG time series and additional topography-based procedures. Channels corresponding to electrodes that were replaced by stimulation electrodes were also excluded. Artifact-contaminated segments were manually identified and removed following visual inspection of the EEG signal. Analyses were restricted to the central 185 scalp EEG channels, excluding neck and face electrodes. Any channels within this subset that were previously removed were interpolated using spherical spline interpolation before average referencing.

Power spectral density was calculated with Welch’s method using 6-second Hanning windows with 90% overlap. Power within a frequency band was defined as the average power across the central 185 scalp EEG channels. EEG power bands were defined as follows:

- Delta: 0.5-4 Hz
- Theta: 4-8 Hz
- Sigma: 11-16 Hz
- Beta: 16-25 Hz
- Gamma: 25-40 Hz

### MRI data acquisition

A 3 Tesla MAGNUS (Microstructure Anatomy Gradient for Neuroimaging with Ultrafast Scanning, GE Healthcare) head-only scanner was used to acquire MRI data. T1- and T2-weighted sequences captured brain structural images with the following parameters:

- T1:

`o` sequence = MP-RAGE (Magnetization-Prepared Rapid Gradient-Echo)
`o` repetition time (TR) = 2000 ms
`o` echo time (TE) = 3 ms
`o` field of view (FOV) = 256 x 256 mm²
`o` inversion time (TI) = 1100 ms
`o` flip angle = 8 degrees
`o` number of slices = 240
`o` resolution = 0.8 mm x 0.8 mm x 0.8 mm
`o` matrix size = 320 x 320 pixels
`o` acquisition time = 4 minutes.

- T2:

`o` sequence = CUBE-T2
`o` TR = 2500 ms
`o` TE = 90 ms
`o` FOV = 256 x 256 mm²
`o` echo train length (ETL) = 120
`o` number of slices = 240
`o` resolution = 0.8 mm x 0.8 mm x 0.8 mm
`o` matrix size = 320 x 320 pixels
`o` acquisition time = 4 minutes.

### TES-TI setup

Silver-silver chloride ceramic electrodes (outer diameter: 12.6 mm; inner diameter: 8.0 mm; width: 2.2 mm) were embedded into the hd-EEG net, replacing standard recording electrodes, and connected to the TI Brain Stimulator for Research (TIBS-R v3.0, TI Solutions AG, Switzerland) for stimulation delivery. Electrode-skin impedance of the stimulating electrodes was continuously monitored and maintained between 4 and 6 kΩ throughout the experiment.

TES-TI was administered using a carrier frequency of 15 kHz. In a subset of participants, balanced across conditions (TES^15k^^Hz^-TI^Peak^ = 3, TES^15k^^Hz^-TI^10Hz^ = 2, TES^15k^^Hz^ = 2), a multipolar montage was employed, i.e., a second set of stimulating electrodes was used to stimulate with the same difference frequency. When a multipolar montage was employed, a carrier frequency of 17 kHz was selected for the second pair of stimulation channels^29^. Three stimulation protocols were tested: (1) TES-TI with a 10 Hz difference frequency (TES^15k^^Hz^-TI^10Hz^), (2) TES-TI with a difference frequency matched to the participant’s individual sigma peak (TES^15k^^Hz^-TI^Peak^), and (3) TES without a difference frequency, that is, only carrier frequency (TES^15k^^Hz^). As a further control condition for the deepening of sleep during the nap, two consecutive 3-minute intervals of N2 sleep were selected from the baseline nap and compared (No Stimulation condition). To make the No Stimulation condition comparable to the stimulation protocols, N2 consecutive 3-minute intervals were randomly selected from different parts of the nap, choosing them at varying time distances from sleep onset. Their number approximately matched the number of stimulation intervals analyzed in the other stimulation protocols with the largest available datasets. All stimulation protocols employed sinusoidal waveforms and, to avoid stimulation effects related to rapid exposure switching^23^, were delivered with a 15-second ramp-up and 15-second ramp-down periods. Stimulation intensity was set to the highest level that did not produce any scalp sensation, up to a maximum of 8 mA peak-to-peak, in line with parameters found safe and effective by previous studies^26^. Stimulation intensity ranged 4-8 mA, with all stimulating electrodes equally delivering the same amount of current for both the kHz carrier and difference frequency stimulation protocols.

Hardware filters placed between the stimulator output and the EEG amplifier eliminated electrical noise from the stimulator, enabling reliable EEG recording during stimulation. A full description of the setup can be found in Schaeffer et al., 2025^30^.

### Optimization of stimulating electrodes placement

Electrode placement was individualized for each participant using TI-Toolbox^31^, an open-source platform for temporal interference research. The toolbox comprises four modules: preprocessing, optimization, simulation, and analysis.

1. Preprocessing: Individualized head models were generated from structural MRI data. DICOM images were first converted to NIfTI volumes and subsequently transformed into meshed volumes conductors using the *charm* function. Tissue-specific conductivity values were assigned to the different tissues, assuming isotropic conductivities throughout the head model. As part of the preprocessing pipeline, the GSN-HydroCel-256 net was automatically co-registered to each individual head model using a non-linear registration from MNI space.
2. Optimization: The optimal electrode montage for targeting both thalami was determined using genetic and/or local exhaustive search algorithms introduced by Haber and colleagues^31^. Stimulation electrodes placement was optimized to maximize the temporal interference electric field at target and minimize it outside the ROI.
3. Simulation: Electric field distributions and TI patterns were computed for the montage(s) identified in Step 2. Electric field values within gray matter were extracted and visualized on both the participant’s native T1-weighted MRI and the standardized MNI152 atlas.
4. Analysis: Intensity and focality metrics were calculated for the thalami. Intensity was defined as the mean TI field modulation magnitude within the ROI, and focality as the ratio of the mean TI field modulation magnitude in the ROI to that in the remaining gray matter. For each participant, the montage selected provided the best trade-off between intensity and focality; specifically, TI field modulation magnitude was maximized at target and minimized off-target.

### Sleep spindle detection

Sleep spindles were detected using the open-source Python library Yet Another Spindle Algorithm (YASA)^32^, which implements an algorithm inspired by the method originally described by Ferrarelli et al.^10^ and Lacourse et al.^33^. EEG data were first bandpass-filtered within the spindle frequency range (Sigma band: 11–16 Hz). Spindle detection was then based on three complementary features:

- Root Mean Square (RMS): the RMS of the filtered signal was computed using a sliding window of 300 ms with a 100 ms step size;
- Band-Specific Correlation: a moving correlation was calculated between the broadband EEG signal (1-30 Hz) and the spindle-band filtered signal, using the same windowing parameters;
- Relative Spectral Power: the proportion of power in the sigma band relative to the total broadband signal power was estimated using Short-Time Fourier Transform with 2-second Hanning windows and 200 ms overlap.

A spindle was detected when the following criteria were simultaneously met:

- the RMS exceeded the mean RMS plus 1.5 times the RMS of successive differences in windows;
- the moving correlation exceeded 0.65;
- the relative sigma power was greater than 0.2.

Events with durations shorter than 0.3 seconds or longer than 2.5 seconds were excluded, regardless of whether they met the detection criteria. Only spindles detected in at least 2 channels were included, while events occurring within 500 ms of each other were considered as the same event.

To comprehensively capture the full spectrum of spindle activity, spindle detection was separately performed across two frequency ranges: 11-13.5Hz and 13-16Hz. The resulting spindle events were subsequently merged, and in cases of overlap, only the longer spindle was used. Integrated spindle activity (ISA) was computed by integrating the absolute amplitude in the sigma band range of each detected spindle across all electrodes within a time interval. It was selected as a primary outcome because it provides a unified measure of multiple spindle characteristics, i.e., amplitude, density, and duration, thereby offering a comprehensive representation of spindle activity within the interval of interest. ISA was computed for both the STIM and PRE normalizing the resulting value by the duration of the interval^10^.

### Statistical analysis

Quantitative variables were summarized using the median (x̃) and interquartile range, while categorical variables were described using frequencies and percentages. The Kruskal-Wallis test was performed to assess potential differences in age and sleep-related metrics across experimental conditions. Fisher’s exact test was used to evaluate differences in gender, race, and ethnicity across stimulation protocols. The Wilcoxon signed-rank test was employed to compare: average band power and ISA during stimulation (STIM) and the preceding three-minute (PRE) (paired); and intensity and focality of the TI field modulation magnitude in the thalami of participants in TES^15k^^Hz^-TI^Peak^ and TES^15k^^Hz^-TI^10Hz^ protocol groups. For topographical comparisons, a cluster-based nonparametric statistical approach was used^34^, based on the method described in Fattinger et al.^35^. Mixed-effects linear regression models were fitted to compare the increase (STIM-PRE) in sigma power and ISA in the TES^15k^^Hz^-TI^10Hz^ protocol compared to the TES^15k^^Hz^-TI^Peak^, TES^15k^^Hz^, and No Stimulation experiment conditions; and to compare the TES^15k^^Hz^-TI^10Hz^ protocol compared to the TES^15k^^Hz^-TI^Peak^ adjusting for the TI field modulation magnitude in the thalami. Analyses were performed using R version 4.4.1, MATLAB 2024a, and Python 3.10.6. All statistical tests were two-sided, with a significance threshold set at p < 0.05.

## Results

### Descriptive statistics of the sample

A total of 48 participants were enrolled in the study, of whom 2 were excluded from all analyses due to the absence of analyzable N2 intervals in any experimental condition. After the initial visit, during which hdEEG data were collected during a nap, participants returned for two additional sessions in which TES-TI targeting the thalamus bilaterally and TES protocols were delivered during N2 sleep in random order (Figure 1).

**Figure 1.**
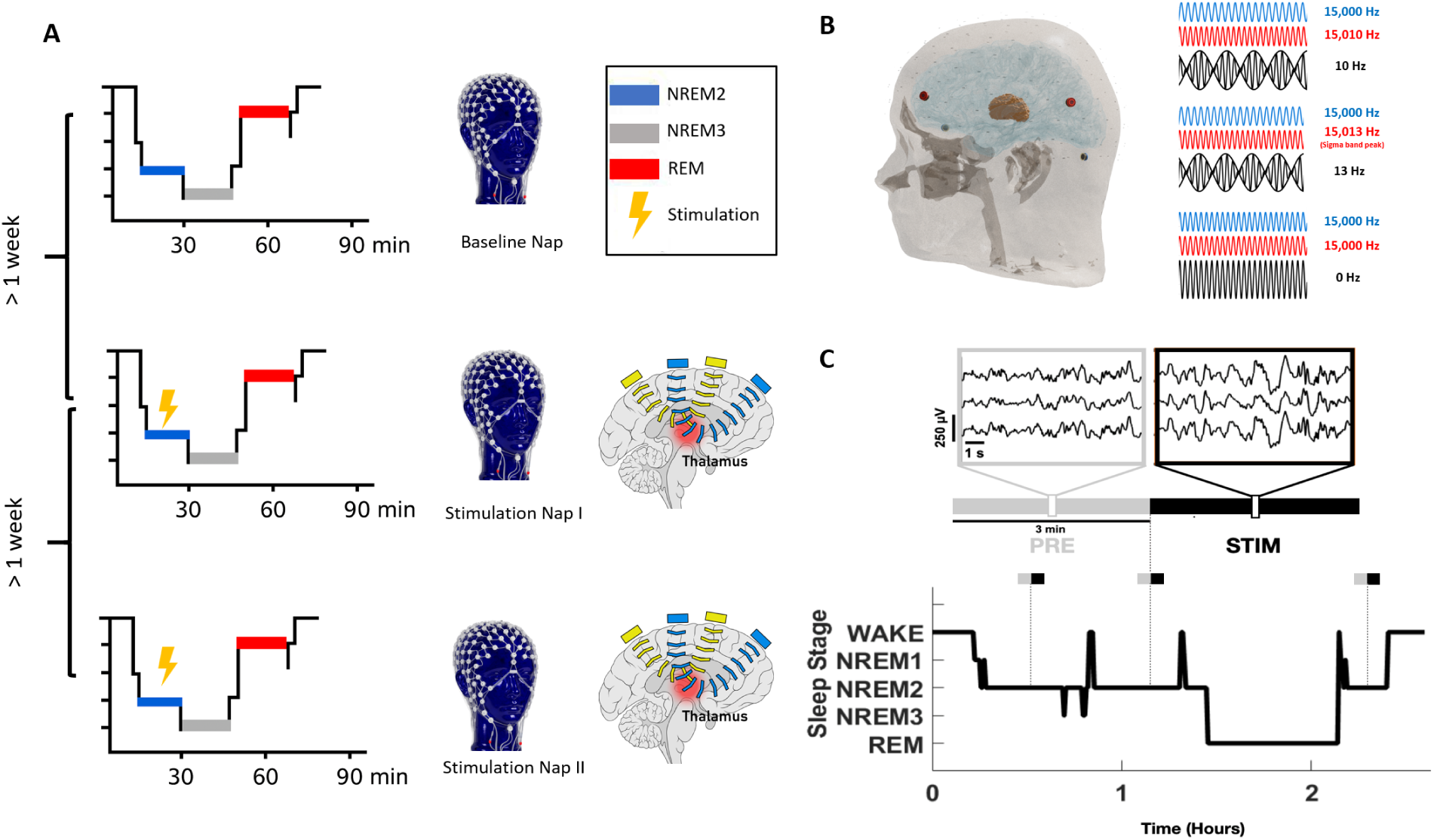
Experiment design. **A**, During this first viit, high-density EEG was recorded without stimulation (Baseline Nap). Two additional visits, scheduled at least one week apart, followed the same procedure but included TES-TI bilaterally targeting the thalamus and TES stimulation protocols during N2 sleep (stimulation Nap I and II **B**, Representative stimulating electrodes’ location for thalamic targeting and a schematic representation of the stimulation frequencies. **C**, Example of stimulation protocols delivered during a Stimulation Nap. Only protocols where both STIM and PRE occurred entirely within N2 sleep were included in the analysis.

Stimulation electrode placement was optimized individually for each participant using head models derived from their T1 and T2-weighted MRI scans (Figure 2A).

**Figure 2.**
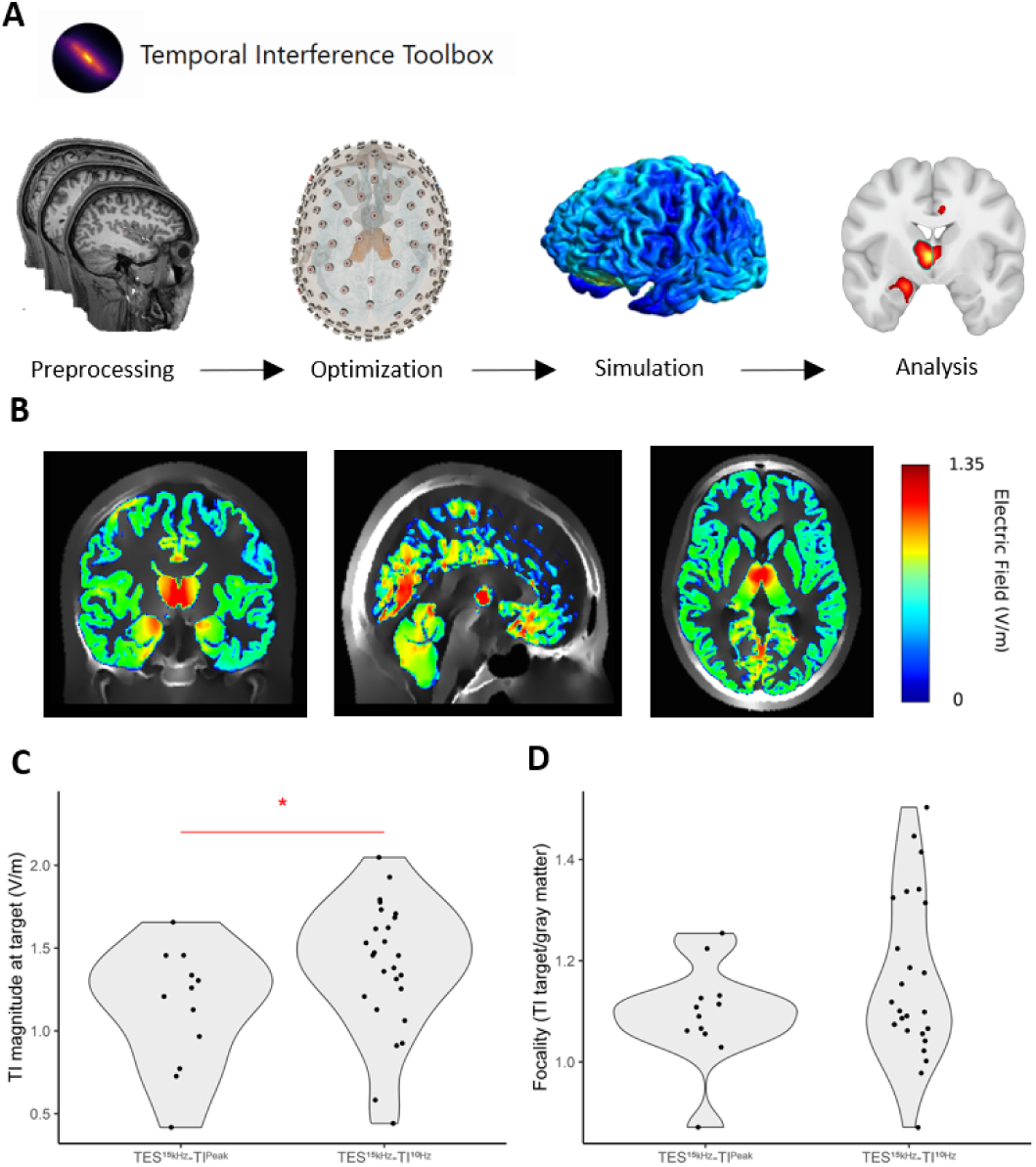
Individual optimization of stimulating electrode placement. **A**, individualized head models were generated from T1- and T2-weighted MRI scans (Preprocessing). Electrode placement was optimized to maximize the temporal interference electric field in the thalami and minimize it outside the region of interest (Optimization). Temporal Interference (Tl) field modulation magnitude distribution across gray matter was visualized to confirm adequate stimulation of the target (Simulation). Intensity and focality of electric field were quantified for each participant (Analysis). **B,** distribution of the simulated Tl field modulation magnitude distribution within the gray matter of a participant of the study, visualized in MNI space. Average Tl field modulation magnitude distribution in the thalami **(C)** and focality of the stimulation **(D)** under two conditions; TES^15k^^Hz^-TI^10Hz^and TES^15k4z^-TI^Peak^ frequency. No significant difference in the focality between the two stimulation protocols was found (Wilcoxon test, p = 0.43), while TES^15k^^Hz^-TI^10Hz^ protocol was associated to higher Tl field modulation magnitude compared to the TES15kHz-TIPeak (Wilcoxon test, p = 0.044).

Only stimulation intervals that were entirely included within the N2 sleep stage and preceded by 3 minutes of N2 sleep were included in the final analysis. As a result, not all participants contributed data to every stimulation protocol. The final distribution was as follows: 25 participants for the TES^15k^^Hz^-TI^10Hz^ protocol, 12 for the TES^15k^^Hz^-TI^Peak^ protocol, and 26 for the TES^15k^^Hz^ protocol. Similarly, 26 participants were included in the baseline condition.

The median of the 46 participants finally included was 24.0 years ± 9.5 ys, and 58.7% of them were female. No significant differences were observed across experiment conditions in demographic variables. Among the nap-related metrics, only sleep onset latency showed a significant difference across conditions, with longer latencies observed during the baseline visit. Results are fully displayed in Table 1.

**Table 1.**
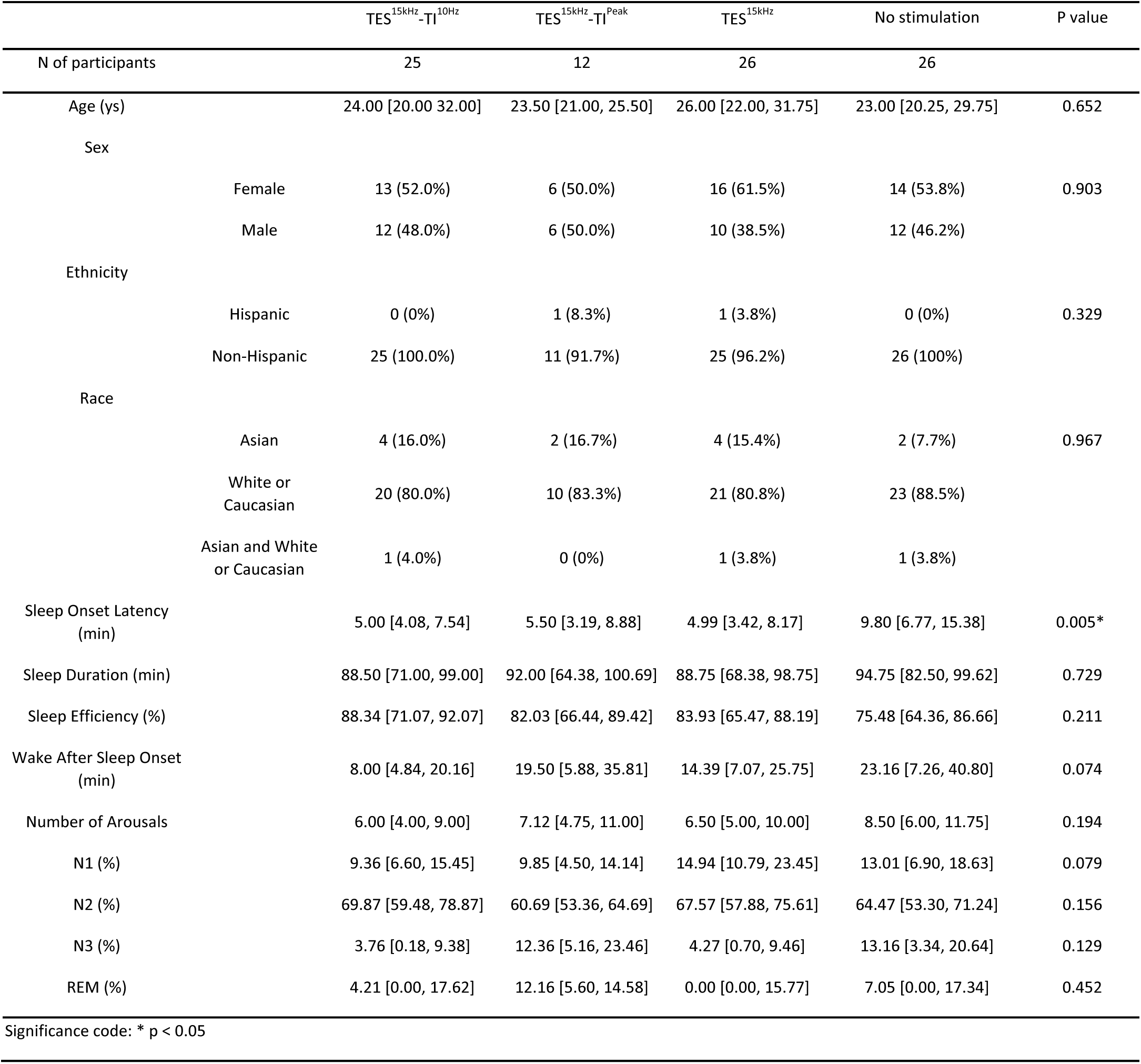
Descriptive statistics of the sample (median [IQR]). Significant differences across experiment conditions were found in sleep onset latency. The variable showed higher values during the “No stimulation” condition, possibly reflecting a “first nap effect”.

### TES-TI can accurately target the thalamus

Figure 2B illustrates the spatial distribution of the simulated TI field modulation magnitude within a participant’s gray matter. The maximum field intensity was localized along the midline, encompassing both the left and right thalami. Within the target regions (i.e., bilateral thalami), the median TI field modulation magnitude across all participants was 1.35 ± 0.47 V/m, and the median focality was 1.10 ± 0.15. These values indicate that, during stimulation, the thalamus was exposed to an electric field of greater strength than the surrounding brain regions and of sufficient intensity to potentially elicit a physiological response.

To evaluate the consistency of stimulation efficacy across stimulation protocols, we compared the mean TI field modulation magnitude at the target and focality between the TES^15k^^Hz^-TI^10Hz^ and TES^15k^^Hz^-TI^Peak^ protocol groups. No significant differences were observed in terms of focality (p = 0.43), while TI field modulation magnitude was significantly higher in the TES^15k^^Hz^-TI^10Hz^ group compared to the TES^15k^^Hz^-TI^Peak^ group (p = 0.044) (Figures 2C–D).

### TES^15kHz^-TI^10Hz^ enhanced spindle activity during stimulation (STIM) compared to pre-stimulation activity (PRE)

To test the hypothesis that TES-TI targeting the thalamus can enhance sleep spindle activity, we first examined sigma band power during STIM compared to PRE. As shown in Figure 3A, the TES^15k^^Hz^–TI^10Hz^ protocol was associated with an increase in sigma band power (Δx̃_STIM–PRE_ = 0.46 ± 1.05 log₁₀ µV², p = 0.0042, roughly corresponding to a threefold increase in power). This was not the case for any of the other experiment conditions, including TES^15k^^Hz^-TI^Peak^ and the TES^15k^^Hz^ protocols (Figure 3A). The latter was instead associated with a significant decrease in sigma power (Δx̃_STIM–PRE_ = -0.32 ± 0.87 log₁₀ µV², p = 0.041, which corresponds approximately to a 50% reduction in power). The No Stimulation condition showed no significant change (Figure 3A).

**Figure 3.**
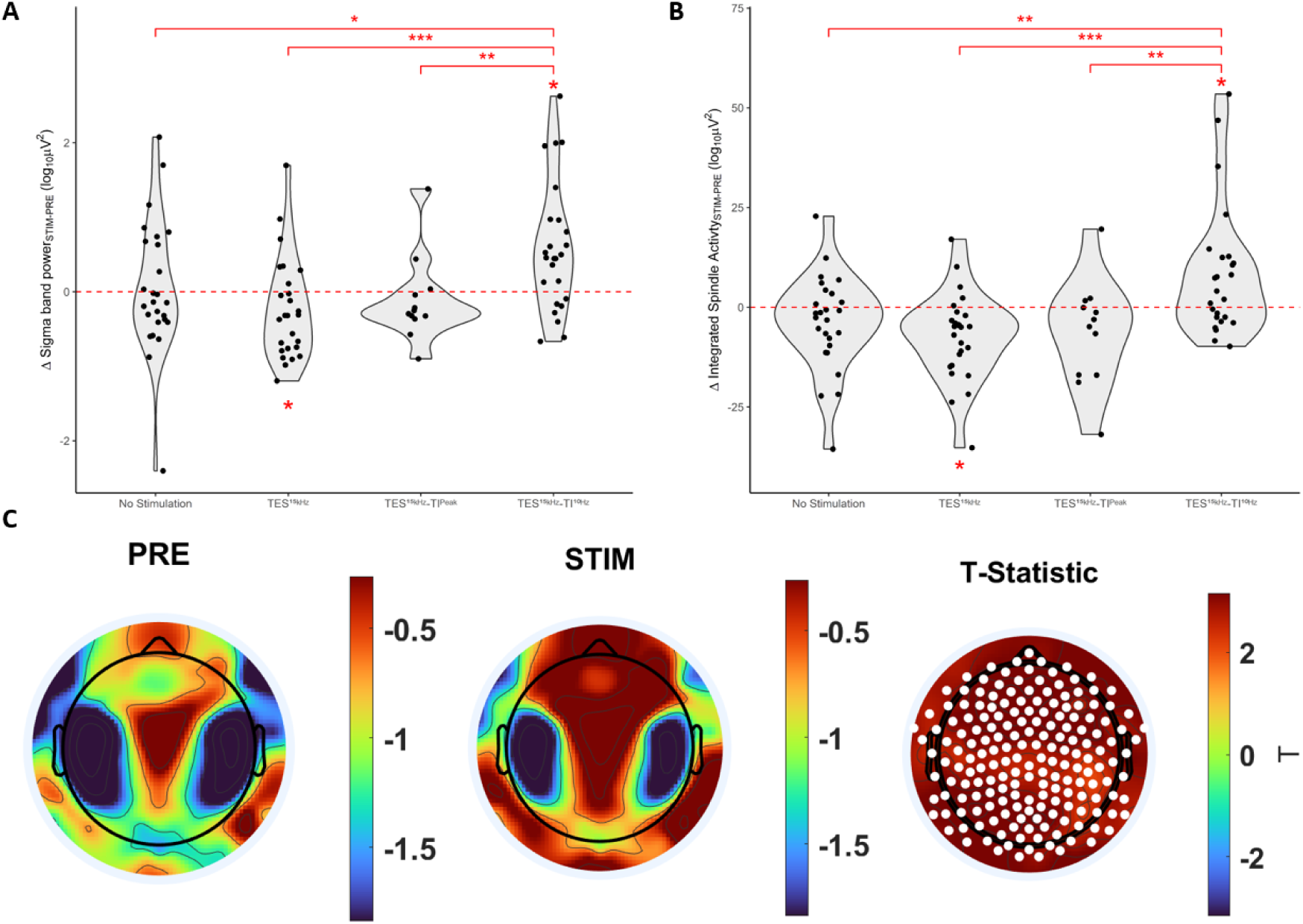
TES^15k^^Hz^-TI^10Hz^ stimulation enhances spindle activity. Distribution of the difference between the stimulation period (STIM) and the preceding three minutes (PRE) in sigma power (11-16Hz) **(A)** and integrated spindle activity (ISA) (B) across the stimulation protocols: TES^15k^^Hz^, i.e., kHz carrier frequency delivered without difference frequency; TES^15k^^Hz^- T|PMk protocol, i.e,, difference frequency matching individual participants’ peak in the sigma range; TES^l5kHz^-TI^10Hz^ protocol, i.e., difference frequency of 10 Hz. Each data point in the plot corresponds to one participant. Red asterisks indicate significance (paired Wilcoxon test for within-subject comparisons, mixed-effects linear regression models for between-subject comparisons; *p<0.05, **p<0.01, *“*p<0.001). **C,** topographical distribution of sigma power during PRE and STIM for the TES^15f^^cHz^-TI^10Hz^stimulation protocol, along with the T-statistic <u>topopjot</u> of electrode comparisons (duster-based permutation). Electrodes showing significant increases after cluster correction are marked in white.

We then examined the topography of the increase in sigma band power in the TES^15k^^Hz^-TI^10Hz^ protocol. The cluster-based permutation analysis conducted revealed a broad, significant positive cluster including all the central 185 EEG electrodes, showing increased sigma power across the entire scalp (average p-alue of electrodes surviving cluster correction = 0.008; Figure 3C). No significant cluster in the sigma band was identified in the other conditions.

Finally, we investigated changes in ISA, a unified measure of multiple spindle features. In the TES^15k^^Hz^-TI^10Hz^ protocol, ISA significantly increased during stimulation compared to the pre-stimulation period (Δx̃_STIM–PRE_ = 4.064 ± 15.00 µV/s, p = 0.030). Consistently with the results described for sigma power, no change was observed in the TES^15k^^Hz^-TI^Peak^ protocol ^(^Δx̃_STIM–PRE_ = –3.96 µV/s ± 17.31, p = 0.11) and during the baseline nap (Δx̃_INT2–INT1_ = -2.049 µV/s ± 12.03, p = 0.13), while the TES^15k^^Hz^ protocol was associated with a significant decrease in ISA (Δx̃_STIM–PRE_ = –4.89 µV/s ± 11.50, p = 0.0014) (Figure 3B).

Linear mixed effects models confirmed the significant increase in sigma band power and spindle features observed between STIM and PRE in the TES^15k^^Hz^-TI^10Hz^ protocol. We fit two mixed-effect models to compare the impact of the 10 Hz difference stimulation, used as the reference for comparisons, with the other experiment conditions. One model used STIM-PRE differences in sigma band power and the other model used ISA as dependent variables. Stimulation protocol was included as a fixed effect, while participant ID was modeled as a random effect to account for repeated measures across conditions. The analysis revealed that the increase in both sigma band power and ISA from PRE to STIM was significantly greater for TES^15k^^Hz^-TI^10Hz^ compared to the other experiment conditions. Results of the linear mixed effects models are presented in Table 2.

**Table 2.** Mixed-effects models were fitted using two outcomes: the increase in sigma power and the integrated spindle activity (ISA) during the stimulation period (STIM) compared to the preceding 3-minute interval (PRE). These models compared the TES^15k^^Hz^-TI^10Hz^ protocol with the other experiment conditions.

| | $\Delta$ Sigma band power ($\log_{10} \mu V^2$) | | | $\Delta$ Integrated spindle activity ($\mu V/s$) | | |
| --- | --- | --- | --- | --- | --- | --- |
| | $\beta$ | Std. Error | p | $\beta$ | Std. Error | p |
| TES<sup>15kHz-TI<sup>10Hz</sup></sup> | 0.58 | 0.15 | 0.00016*** | 8.28 | 2.57 | 0.0013** |
| TES<sup>15kHz-TI<sup>Peak</sup></sup> | -0.70 | 0.27 | 0.0095** | -14.70 | 4.52 | 0.0011** |
| TES<sup>15kHz</sup> | -0.83 | 0.22 | 0.00013*** | -15.66 | 3.60 | 0.000014*** |
| No Stimulation | -0.53 | 0.22 | 0.013* | -12.11 | 3.60 | 0.00078*** |

Significance code: \* $p < 0.05$ , \*\* $p < 0.01$ , \*\*\* $p < 0.001$

To ensure that the observed associations between TES^15k^^Hz^-TI^10Hz^ stimulation and increases in sigma power and ISA were not confounded by the significantly greater TI field modulation magnitude in the ROI in this group compared with the participants who received the TES^15k^^Hz^-TI^Peak^ stimulation, we fitted two additional mixed-effects models. These models directly compared the TES^15k^^Hz^-TI^10Hz^ and TES^15k^^Hz^-TI^Peak^ groups in terms of STIM–PRE differences in sigma power and ISA, while adjusting for TI field modulation magnitude. The between-group differences remained significant, and TI field modulation magnitude did not predict either outcome. These findings indicate that the greater efficacy of the 10 Hz difference frequency, relative to a difference frequency matched to the individual sigma peak, cannot be explained by differences in TI field modulation magnitude. Results are presented in Table 3.

**Table 3.** Mixed-effects models were fitted using two outcomes: the increase in sigma power and the integrated spindle activity (ISA) during the stimulation period (STIM) compared to the preceding 3-minute interval (PRE). These models compared the TES^15k^^Hz^-TI^10Hz^ protocol the TES^15k^^Hz^-TI^Peak^ protocol, adjusting for the TI field modulation magnitude in the thalami. TES^15k^^Hz^-TI^Peak^ protocol row was left blank as it represents the reference for comparison.

| | $\Delta$ Sigma band power ($\log_{10} \mu V^2$) | | | $\Delta$ Integrated spindle activity ($\mu V/s$) | | |
| --- | --- | --- | --- | --- | --- | --- |
| | $\beta$ | Std. Error | p | $\beta$ | Std. Error | p |
| Intercept | -0.034 | 0.45 | 0.94 | -4.75 | -0.54 | 0.59 |
| TES<sup>15kHz</sup>-TI<sup>Peak</sup> |  |  |  |  |  |  |
| TES<sup>15kHz</sup>-TI<sup>10Hz</sup> | 0.72 | 0.28 | 0.011* | 15.10 | 5.57 | 0.0067** |
| TI modulation magnitude | -0.075 | 0.34 | 0.83 | -1.46 | 6.75 | 0.83 |

Significance code: \* $p < 0.05$ , \*\* $p < 0.01$ , \*\*\* $p < 0.001$

### Low-frequency EEG power increased and high-frequency power decreased in all protocols between PRE and STIM epochs, reflecting increasing sleep depth during N2

Our analyses are based on one or two stimulations per participant, administered exclusively during the N2 sleep stage. Owing to the deepening of sleep within the nap, N2 EEG spectral power is expected to change over time regardless of the stimulation protocol employed. This factor can be averaged out in overnight studies where multiple stimulations can be delivered at different points in a longer sleep episode^30^. To tease apart spectral changes attributable to the spontaneous evolution of N2 sleep from those related to TES-TI stimulation, we compared two consecutive 3-minute intervals of N2 sleep extracted from the baseline nap, when no stimulation was delivered.

Consistent with the evolution of N2 sleep, for the No Stimulation condition, the second 3-minute interval exhibited a significant increase in delta band power (Δx̃_INT2-INT1_ = 0.85 ± 0.75 log₁₀ µV², p = 0.0007) and theta band power (Δx̃_INT2-INT1_ = 0.27 ± 0.78 log₁₀ µV², p = 0.0020), along with significant decreases in beta (Δx̃_INT2-INT1_ = -0.44 ± 0.71 log₁₀ µV², p = 0.00022) and gamma band power (Δx̃_INT2-INT1_ = -0.46 ± 0.48 log₁₀ µV², p = 0.012). No significant change was observed in the sigma band (Δx̃_INT2-INT1_ = -0.040 ± 1.05 log₁₀ µV², p = 0.69). Comparable spectral changes between STIM and PRE were observed in the TES^15k^^Hz^-TI^Peak^ protocol without concomitant changes in the sigma band power (Δx̃_STIM-PRE_ = -0.27 ± 0.34 log₁₀ µV², p = 0.20); and in the TES^15k^^Hz^ protocol associated to a significant decrease in the sigma band power (Δx̃_STIM-PRE_ = -0.32 ± 0.87 log₁₀ µV², p = 0.041). Thus, TES^15k^^Hz^-TI^10Hz^ was the only stimulation protocol to show increases in sigma power, in addition to the expected changes associated with deepening of sleep. Results are reported in Table 4 and displayed in Figure 4.

**Figure 4.**
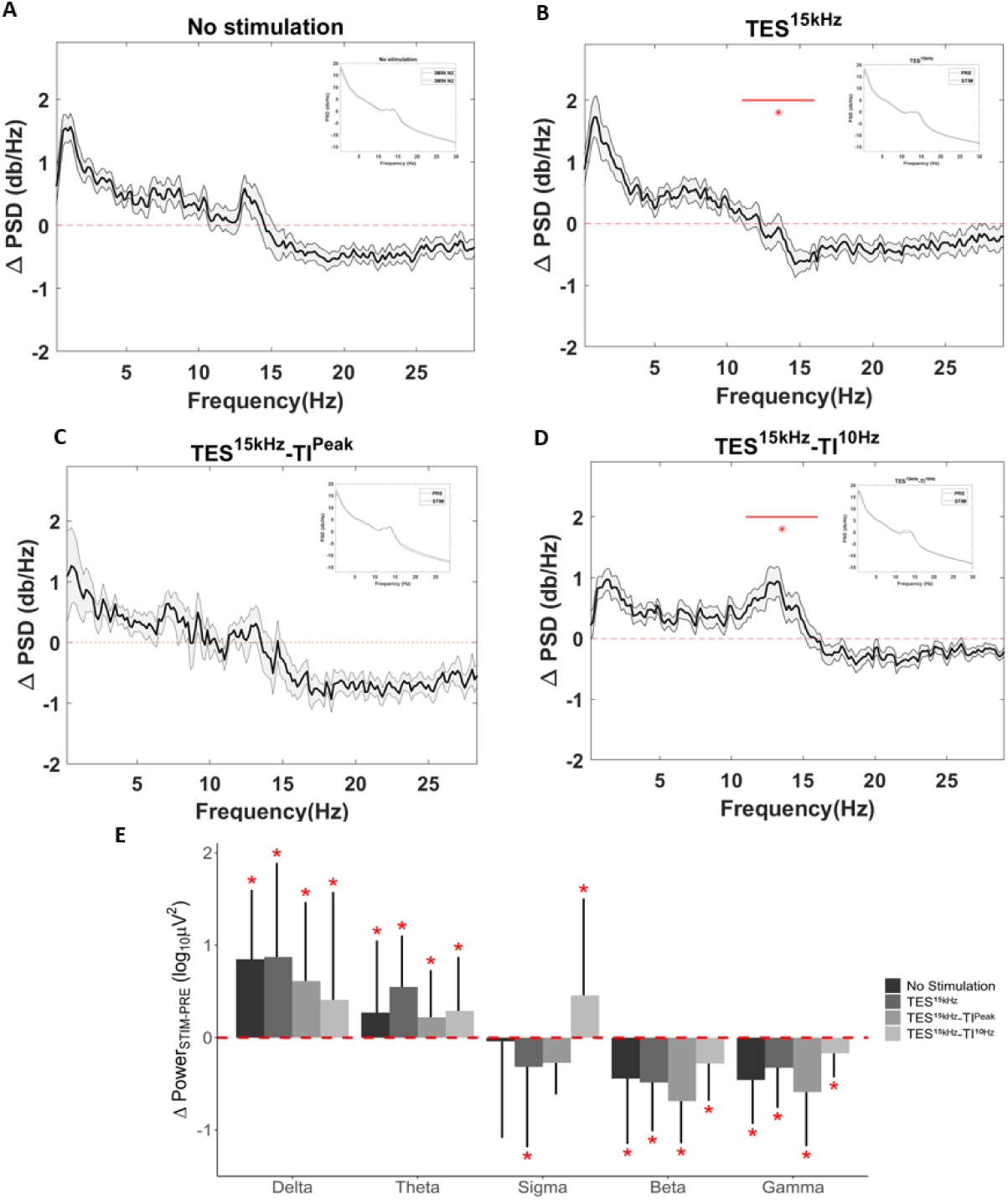
Changes in EEG spectral power changes across conditions. **A**, difference in power spectra between two consecutive 3-minute intervals of N2 during the baseline nap. **B,** difference in power spectra between SUM and PRE in the TES^15k^^Hz^ protocol, i.e., kHz carrier frequency delivered without envelope frequency. **C,** difference in power spectra between STIM and PRE in the TES^15k^^Hz^-TI^Peak^ protocol, i.e., difference frequency matching individual participants’ peak in the sigma range. **D,** difference in power spectra between STIM and PRE in the TES^15k^^Hz^-TI^10Hz^ protocol, i.e., difference frequency of 10 **Hz.** Average across electrodes, then across participants (dark line) and standard error of the mean difference (shaded light gray areas) are shown in each panel. The power spectra of the two intervals compared are displayed in the top right insert and they all exhibit a typical N2 sleep profile, with a dominant delta peak (slow waves) and a secondary sigma peak (spindles). **E,** changes in the frequency bands across the conditions at hand. Bar plots show the median difference, with error bars indicating interquartile range. Red asterisks indicate significance (paired Wilcoxon test, p <0.05).

**Table 4.** Median and interquartile range of the EEG spectral power differences (log₁₀ µV²) between a 3-minute stimulation interval and the preceding 3 minutes across three conditions: TES-TI with a 10 Hz difference frequency (TES^15k^^Hz^-TI^10Hz^), TES-TI with a difference frequency matching the individual sigma peak (TES^15k^^Hz^-TI^Peak^), kilohertz carrier frequency only (TES^15k^^Hz^); and a no stimulation condition, using two consecutive N2 intervals from the baseline nap.

| | $\text{TES}^{15\text{kHz-TI}^{10\text{Hz}}}$ | | $\text{TES}^{15\text{kHz-TI}^{\text{Peak}}}$ | | $\text{TES}^{15\text{kHz}}$ | | No Stimulation | |
| --- | --- | --- | --- | --- | --- | --- | --- | --- |
| EEG band | $\Delta\tilde{x} \pm \text{IQR}$ | p | $\Delta\tilde{x} \pm \text{IQR}$ | p | $\Delta\tilde{x} \pm \text{IQR}$ | p | $\Delta\tilde{x} \pm \text{IQR}$ | p |
| Delta | $0.41 \pm 1.17$ | 0.0023** | $0.61 \pm 0.85$ | 0.016* | $0.87 \pm 1.02$ | 0.00049*** | $0.85 \pm 0.75$ | 0.00007*** |
| Theta | 0.29 ± 0.59 | 0.00049*** | 0.22 ± 0.51 | 0.016* | 0.55 ± 0.56 | 0.0013** | 0.27 ± 0.78 | 0.0020** |
| Sigma | 0.46 ± 1.05 | 0.0042** | -0.27 ± 0.34 | 0.20 | -0.32 ± 0.87 | 0.041* | -0.040 ± 1.05 | 0.69 |
| Beta | -0.28 ± 0.40 | 0.0049** | -0.69 ± 0.45 | 0.00098*** | -0.49 ± 0.53 | 0.00058*** | -0.44 ± 0.71 | 0.00022*** |
| Gamma | -0.17 ± 0.26 | 0.00073*** | -0.59 ± 0.59 | 0.0049** | -0.33 ± 0.43 | 0.00084*** | -0.46 ± 0.48 | 0.012* |

Significance code: \* p < 0.05, \*\* p < 0.01, \*\*\* p < 0.001

## Discussion

In this study, we demonstrated that TES-TI targeting the thalamus can effectively enhance spindle activity during a daytime sleep episode. We compared three minutes of stimulation (STIM) to the preceding three minutes (PRE) across three stimulation protocols: TES^15k^^Hz^-TI^10Hz^ (15 kHz carriers with a 10 Hz difference frequency); TES^15k^^Hz^-TI^Peak^ (15 kHz carriers with a difference frequency matched to each participant’s peak within the sigma power band); TES^15k^^Hz^ (15 kHz carriers with 0 Hz difference frequency). Data from an additional no stimulation control (two consecutive 3-minute N2 sleep intervals from baseline nap) were also analyzed to isolate changes in the EEG power spectrum attributable to increasing sleep depth during the nap. The results show that the TES^15k^^Hz^-TI^10Hz^ protocol significantly increased sigma power during STIM compared to PRE, while none of the other protocols did. Cluster-based permutation analysis localized this increase across the entire scalp. The increase in sigma power was associated with a significant increase in ISA, defined as the integrated absolute amplitude of the sleep spindles detected across all electrodes in a time interval of interest. Linear mixed effects models showed that the increase in both sigma band power and ISA during STIM compared to PRE was greater in the TES^15k^^Hz^-TI^10Hz^ compared to the TES^15k^^Hz^ protocol, the TES^15k^^Hz^-TI^Peak^ stimulation and the No Stimulation condition.

Our experimental design employs a limited set of stimulations administered during N2 sleep within brief daytime nap episodes. This approach offers the simplicity necessary to assess the feasibility of a novel technique. However, it is susceptible to be confounded by shifts in the EEG power spectra associated with the progressive deepening of sleep within this stage in the course of a nap, a source of variability that would have been mitigated if multiple stimulation intervals at different points of a night sleep episode were averaged^30^. To disentangle possible time-related effects from those genuinely attributable to stimulation, we compared two consecutive 3-minute N2 intervals from the baseline nap, during which no stimulation was applied, to mirror pre-stimulation and stimulation intervals. In the second interval relative to the first, we observed an increase in delta and theta power and a decrease in beta and gamma power. Similar patterns, i.e., an increase in low-frequency power and a decrease in high-frequency power, were consistently observed across all experimental conditions. These spectral shifts are characteristic of the increasing sleep depth during N2 sleep^36^, which often represents a transition state between “light” N1 sleep and “deep” N3 sleep^37^. This finding confirms that TES-TI stimulation preserves physiological sleep architecture while it can selectively act on its target, namely spindle activity. This conclusion is also supported by the fact that most nap-related metrics (i.e., sleep duration, sleep efficiency, wake after sleep onset, number of arousals, sleep stages distribution) did not differ across conditions, indicating that TES-TI administration during sleep neither induced awakenings nor disrupted sleep architecture. The only significant difference in sleep parameters was observed for sleep onset latency, which was longer in the No Stimulation condition compared with all other conditions. The No Stimulation intervals were selected from the baseline nap, always conducted during the participant’s first study visit. This increase in sleep onset latency may therefore reflect a “first-nap effect”^38^.

Unlike the TES^15k^^Hz^-TI^10Hz^ protocol, tailoring the difference frequency to the peak spindle frequency for each individual participant surprisingly did not have any significant effect on spindle activity. Mixed-effects linear regression models that included TI modulation magnitude in the thalamus as a covariate allowed us to rule out the possibility that differences in targeting quality between participants receiving TES^15k^^Hz^-TI^10Hz^ and the TES^15k^^Hz^-TI^Peak^ protocol accounted for the observed effect. Individualized stimulation frequencies offer potential advantages^39–41^, even though these have not been borne out by studies that directly compared standardized and individualized frequencies using tACS, either during sleep^42^ or during wakefulness^43^. In particular, it has been reported that tACS applied to the occipital cortex at a frequency slightly below the individual alpha peak maximized post-stimulation alpha power in the targeted region^43^. Moreover, recent electrophysiological investigations in rhesus monkeys using single-unit recordings indicate that TES-TI can effectively disrupt synchronized neuronal activity^44^. Thus, TES-TI applied at a difference frequency closely matching the optimal synchronization frequency of target neurons may be less effective or even counterproductive, whereas at nearby frequencies it may help entrain neuronal activity. The TES^15^ ^kHz^ protocol was instead associated with a significant decrease of both sigma power and ISA. This functions as an important active control, as kHz frequency stimulation has been associated with a variety of subthreshold and suprathreshold neural effects ranging from facilitation to desynchronization of neural firing and even conduction block^45^. Moreover, the absence of a difference frequency produces continuous constructive interference at the target site, yielding the highest root mean square current value of all stimulation protocols. While on one side this result shows that the presence of an amplitude-modulated signal is necessary to induce the intended effect, on the other it suggests that the kHz-frequency sine waves used as carriers can influence neural activity in ways that unexpectedly interfere with the desired outcome^46^. Although previous work has suggested that increasing carrier frequency minimizes off-target effects of TES-TI due to kHz stimulation^47^, further research is warranted to elucidate the mechanisms by which it can affect neural activity in the context of both amplitude-modulated and non-modulated stimulation.

More generally, the mechanisms through which TES-TI acts inside the brain are still unclear. While initial models proposed selective neuronal sensitivity to the low frequency envelope via membrane lowpass filtering^23^, subsequent work indicates that additional mechanisms, such as network level adaptation^48^ or nonlinear single cell frequency mixing mediated by voltage gated ion channels^49^, are required, although these effects have been demonstrated only at field strengths exceeding those achievable in humans. Since the peak frequency for most subjects is considerably higher than 10Hz, it will be important to determine whether TES-TI in the fast spindle range, but outside the individual peak frequency, may also be effective.

The feasibility of using TES-TI to enhance specific sleep features without compromising sleep integrity gives this technique advantages against other approaches explored to increase spindle activity. Z-drugs and benzodiazepines have been associated with a reduction in slow wave sleep^18^, while auditory stimulation can involuntarily cause awakening due to the high sensitivity of the thalamo-cortical complex to acoustic stimuli during sleep^21^. As shown by Lustenberger and colleagues^50^, tACS applied to the frontal cortex within the sigma frequency range, in phase with spontaneously occurring spindles, can lead to increased post-stimulation sigma power and improve memory consolidation. However, conventional TES has two limitations that TES-TI was able to overcome. First, it prevents simultaneous recording of EEG due to stimulation artifacts, whereas this is possible at the high carrier frequencies of TES-TI thanks to the use of hardware low-pass filters^30^. Second, it can induce scalp discomfort, possibly interfering with sleep maintenance, when trying to achieve an electrical field of intensity sufficient to induce a response in deep brain structures. This makes it unfeasible to elicit spindles by directly targeting the subcortical structures responsible for their generation, i.e., the thalamus^7,51^. The high frequency carrier employed in TES-TI allowed us to employ higher stimulation intensities without eliciting any sensation^25^, because of the increased skin stimulation threshold, and to enhance spindles by directly stimulating the site of their thalamic source deep inside the brain.

### Strengths and limitations

This study provides evidence of effective non-invasive thalamic stimulation in humans. It also represents the first demonstration of TES-TI efficacy in enhancing spindle activity, validated across multiple active and inactive control conditions. Additionally, our findings confirm the feasibility of TES-TI administration during sleep while concurrently recording EEG, paving the way for future studies testing its use in research and clinical settings. These results also support the specificity of TES-TI stimulation protocols with respect to both difference frequency and target region, as our previous study using TES-TI to target the ventromedial prefrontal cortex with a 1 Hz envelope frequency during sleep showed an increase in slow wave, but not spindle activity^30^. We utilized the open-source TI-Toolbox^31^ platform to individualize electrode placement based on participants’ T1- and T2-weighted MRI scans. Montage optimization was conducted to maximize the focality-intensity trade-off, ensuring that the thalamus received a stronger TI field modulation magnitude than adjacent gray matter regions and that the field strength was sufficient to possibly elicit a physiological response. This assumption is based on previous studies indicating that effective TI field modulation magnitude within targeted regions is frequently below the one achieved in this study participants^52–54^. Additionally, we ensured that any observed differences in outcomes could not be attributed to variations in targeting quality of the ROI across stimulation protocols.

This initial study has obvious limitations. Stimulations were performed during an afternoon nap, which limited each experiment to one or two stimulations administered to each participant. The number of subjects is limited, reducing the power to detect weaker effects. Moreover, some subjects also received additional brief (<=300 s) stimulations during REM sleep or during wakefulness, even if these are unlikely to have affected comparisons between STIM and PRE intervals within N2. To the best of our knowledge the after effects of short TES-TI stimulation beyond a 3-minute post-stimulation interval^30^ have not been systematically investigated. Evidence from TES suggests that stimulation periods of more than 5 minutes are required to produce effects that outlasts the stimulation interval^55^. If these findings generalize to TES-TI, carryover effects of REM and wake stimulations are unlikely to overlap with N2 stimulation or the preceding 3 minutes. If they do not, two factors could minimize their potential influence as confounders. First, these stimulations were delivered using different frequencies, different cortical targets, and in different vigilance states (REM or wake), making any possible resulting effects unlikely to resemble those following N2 stimulation targeting the thalamus within or immediately below spindle range. Second, they were randomly distributed across various experiment conditions, reducing the risk of systematically biasing our results. Finally, while these findings serve as proof of concept for the capacity of thalamic TES-TI stimulation to elicit sleep spindles, their replication in studies employing multiple stimulation intervals across a full night of sleep is necessary. Moreover, further investigation is required to assess the potential impact of TES-TI induced spindle enhancement on memory consolidation and, more generally, on human cognition.

## Conclusion

Bilateral thalamic stimulation via TES-TI during sleep stage N2 delivered with a difference frequency of 10 Hz resulted in an enhancement of spindle activity localized to the entire scalp. The use of a stimulation frequency matched with each participant’s peak in the sigma band failed to achieve the same effect. Further research is required to understand the mechanisms of action of TES-TI in the thalamus, the possible cognitive effects of sleep spindles enhancement, and the potential applications of this technique in diseases associated with sleep spindle deficits (e.g., schizophrenia, Alzheimer’s disease, Parkinson’s disease, epilepsy).

## Declaration of generative AI use

During the preparation of this work, the authors used Copilot for text and code editing. After using this tool, the authors reviewed and edited the content as needed and take full responsibility for the content of the published article.

## Data and Code Availability

The data underlying this article will be shared on reasonable request to the corresponding authors. Customized codes used to create the main figures and tables have been made publicly available on GitHub, at https://github.com/sbruno3/TES-TI_spindles.

## Acknowledgements

The TIBS-R investigational devices used in this project were provided by TI Solutions AG, Switzerland, as part of its Early Adopter Program (www.temporalinterference.com).

